# Evaluating the Multimodal Capabilities of Generative AI in Complex Clinical Diagnostics

**DOI:** 10.1101/2023.11.01.23297938

**Authors:** Marc Cicero Schubert, Maximilian Lasotta, Felix Sahm, Wolfgang Wick, Varun Venkataramani

## Abstract

In the rapidly evolving landscape of artificial intelligence (AI) in healthcare, the study explores the diagnostic capabilities of Generative Pre-trained Transformer 4 Vision (GPT-4V) in complex clinical scenarios involving both medical imaging and textual patient data. Conducted over a week in October 2023, the study employed 93 cases from the New England Journal of Medicine’s image challenges. These cases were categorized into four types based on the nature of the imaging data, ranging from radiological scans to pathological slides. GPT-4V’s diagnostic performance was evaluated using multimodal inputs (text and image), text-only, and image-only prompts. The results indicate that GPT-4V’s diagnostic accuracy was highest when provided with multimodal inputs, aligning with the confirmed diagnoses in 80.6% of cases. In contrast, text-only and image-only inputs yielded lower accuracies of 66.7% and 45.2%, respectively (after correcting for random guessing: multimodal: 70.5 %, text only: 54.3 %, image only: 29.3 %). No significant variation was observed in the model’s performance across different types of images or medical specialties. The study substantiates the utility of multimodal AI models like GPT-4V as potential aids in clinical diagnostics. However, the proprietary nature of the model’s training data and architecture warrants further investigation to uncover biases and limitations. Future research should aim to corroborate these findings with real-world clinical data while considering ethical and privacy concerns.

## Introduction

The emergence of advanced artificial intelligence (AI) technologies has given rise to generative models proficient in text-based interactions and, more recently, visual data interpretation ^1^. While these models have shown promise in academic medical evaluations^2,3^, their practical application in real-world clinical scenarios, particularly those involving a combination of textual and visual data, remains unexamined. This investigation focuses on the diagnostic capabilities of the Generative Pre-trained Transformer 4 Vision (GPT-4V)^4^ in intricate clinical cases that involve both medical imaging and patient case histories.

## Methods

The study was conducted between October 18-25 2023. New England Journal of Medicine *Image challenges* from the GPT-4V training cutoff onwards (January 2022) were used (n = 93, eTable 1), which offer medical cases with verified pathological outcomes. We categorized the clinical image challenges into four broad types: 1) clinical images, 2) radiological scans (MRI, CT, X-ray, Sonography), 3) pathological images (histological slides, macroscopic specimens) and 4) images from other medical procedures. For all challenges, to ensure the model uses the image itself for diagnostic analysis, descriptions about the images were removed from the clinical information text. The model was prompted a) multimodally (with both text and images), b) clinical information text only and c) image only. The primary metric was the alignment of GPT-4V’s chosen diagnosis with the verified pathological outcome based on multimodal input compared to text or image only input. Secondary measures were performance differences between above defined image categories as well as differences between medical specialties. Adjustment for random guessing was performed^5^. The Chi-squared test was used to compare performance differences. Statistical evaluations were performed using R (version 4.0.5).

## Results

Among the 93 cases, GPT-4V’s diagnosis corresponded with the confirmed diagnosis in 80.6 % of cases (75 of 93), while it was capable of answering the challenges based on text only in 66.7 % of cases (62 of 93) and based on the images only in 45.2% of cases (42 of 93), (after correcting for random guessing: multimodal: 70.5 %, text only: 54.3 %, image only: 29.3 %). GPT-4V performed significantly better using multimodal input compared to text input only and image input only (p=.046, p<.001). Between radiological images, clinical images or pathological images, performance was similar (Fig 1, p=.91). No significant differences were observed when comparing different specialties (Fig 1, p=.51).

**Figure 1.**
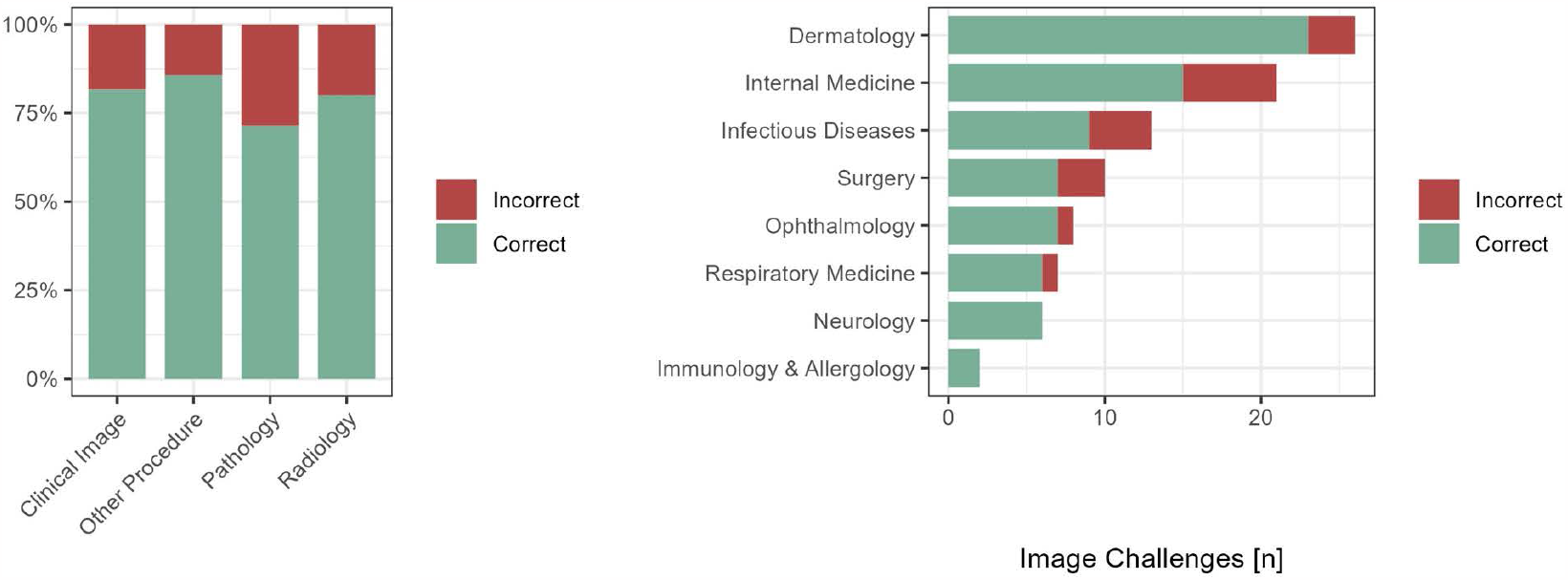
Left: Performance across image categories. Right: Performance across specialties (n = 93 image challenges).

## Discussion

GPT-4V exhibited a high diagnostic accuracy rate of complex multimodal clinical challenges that included images of various categories and clinical data. The investigation shows that GPT-4V is able to increase its diagnostic accuracy by integrating information from both data sources compared to when each data source is used alone. This is analogous to clinical reasoning in practice, where decisions are derived from various information and data sources. The interpretation of clinical images from various sources by GPT-4V itself yielded a low accuracy while it still increased diagnostic accuracy in a multimodal context. With further improvements, multimodal generative AI models like GPT-4V could serve as valuable adjuncts to human diagnostic efforts in the future. However, as training data and its exact architecture are not open-source, further exploration is needed to identify any inherent biases and additional limitations. Given that the image challenges in this study serve primarily as educational tools, future research should aim to validate these findings using real-world clinical data, while adhering to privacy and ethical considerations^6^.

## Data sharing statement

No patient data was used in this study. Image challenges and their solutions are publicly available at https://www.nejm.org/image-challenge. Software for using the Open AI API has been deposited on GitHub at https://github.com/venkataramani-lab/ImageChallenges.

## Acknowledgments

No funding. W.W. is inventor of the patent WO2017020982A1 Agents for use in the treatment of glioma. This patent covers new treatment strategies that all target the formation and function of TMs in glioma.

